# DNA methylation signature of birthweight generalizes to high-risk pregnancies and is independent of genetic, maternal, and obstetric factors: a twin study

**DOI:** 10.64898/2026.08.25.26361321

**Authors:** Mahnoor Sulaiman, Laura Franken, Jip A. Spekman, Sophie G. Groene, Erik W. van Zwet, Arno A.W. Roest, Monique C. Haak, Tom Kuipers, Hailiang Mei, Alexander Neumann, Charlotte Cecil, Bastiaan T. Heijmans

**Author notes:** Shared last author (Charlotte Cecil, Bastiaan T. Heijmans).

## Abstract

**Background:** DNA methylation patterns in cord blood are robustly associated with birthweight in the general population. However, it remains unknown whether these associations extend to clinically relevant populations, such as preterm neonates or those born small for gestational age, and whether they directly reflect birthweight or are driven indirectly by genetic, familial, maternal, and obstetric factors.

**Methods:** We calculated a birthweight methylation profile score (MPS_BW_) using weights of 835 CpGs previously associated with birthweight in the general population and evaluated its association with birthweight in 67 monochorionic (MC) twin pairs including 134 neonates (97% born preterm) from the Twinlife study. MC twin pairs are identical twins sharing a single placenta, often unequally, which can result in unequal resource distribution and differential fetal growth.

**Results:** We examined the association between within-pair differences in birthweight and MPS_BW_, thereby estimating the association independent of factors shared equally by co-twins. A 500-gram increase in birthweight was associated with a 0.256 SD increase in MPS_BW_ (p<0.005) in this population of preterm neonates. Adjustment for polygenic score for birthweight (PGS_BW_) confirmed that the observed epigenetic associations were not driven by common genetic variation underlying birthweight. Interestingly, a similar effect size (0.226 SD per 500 g birthweight increase; p<0.05) was observed in the within-pair analysis, which controls for all shared influences within a twin pair.

**Conclusion:** DNA methylation is associated with individual differences in birthweight in a high-risk clinical population of MC twins, independent of shared genetic, familial or maternal influences.

## Introduction

Birthweight reflects the interplay between genetic influences and intrauterine conditions during prenatal development. Beyond its role as a marker of prenatal exposures, it is also predictive of both neonatal and long-term health outcomes.^5,15,30,38^ However, the biological mechanisms reflecting birthweight variation and their downstream effects on health outcomes remain incompletely understood.

DNA methylation (DNAm) is a strong candidate to serve as such a marker,^18^ as it may capture the cumulative molecular imprint of fetal growth.^7,24,26^ Consistent with this, epigenome-wide association studies (EWAS) have identified CpG sites robustly associated with birthweight in cord blood.^1,25,35^ In particular, a meta-analysis of 8,825 term neonates within the Pregnancy And Childhood Epigenetics (PACE) consortium identified 914 birthweight-associated CpGs in cord blood.^25^ However, these findings are largely derived from healthy neonates born at term, with unclear relevance to clinical populations. Moreover, the factors underlying the association between birthweight and DNA methylation have not been fully disentangled. It remains unclear to what extent these associations are confounded by genetics,^3,39^ familial environment, maternal and obstetric factors,^17,23^ with genetic effects in particular shown to account for part of the birthweight–DNA methylation signal.^8^

Monochorionic (MC) twin pairs provide a natural experiment to disentangle these sources of variation. Co-twins within an MC twin pair share essentially the same genetic background and many maternal, familial, and obstetric factors. Consequently, differences in birthweight and DNAm between co-twins are less likely to be attributable to these equally shared factors.^13,16^ At the same time, they share a single placenta with vascular connections between the two fetuses. Unequal placental sharing and imbalances in intertwin blood flow can result in differences in nutrient and oxygen supply, leading to differential fetal growth and birthweight differences within MC twin pairs.^20,27^ This setting therefore provides a unique framework to study intrauterine influences while accounting for shared genetic, maternal, familial and obstetric factors.

Within the Twinlife study, we analyzed 67 MC twin pairs (n = 134 neonates) who were commonly born preterm and small-for-gestational-age (SGA) with varying levels of birthweight discordance. We examined whether the association between birthweight and methylation identified in the general population extends to these high-risk neonates and whether the association was independent of genetics and shared familial, maternal, or obstetric factors. To this end, we computed a birthweight methylation profile score (MPS_BW_),^34^ which aggregates birthweight-associated CpGs reported within the PACE consortium meta-analysis^25^ into a single score per neonate^8^ and evaluated its association with birthweight. Specifically, we examined the association in preterm neonates at the unpaired, individual level and compared it with the association observed within twin pairs, finding that MPS_BW_ is associated with birthweight in this high-risk population, independent of genetic liability and shared familial, maternal, and obstetric factors.

## Materials and Methods

### Study Participants

This study is part of the Twin Longitudinal Investigation of Fetal Discordance (Twinlife) study, registered under the International Clinical Trials Registry Platform ID NL7538 at Leiden University Medical Center (LUMC), the Netherlands’ leading referral center for complex MC twin pregnancies^19^. This is a longitudinal cohort study approved by the institutional medical ethical committee (P18.184). Written informed consent was obtained from all parents for blood collection, isolation, and DNA analysis. Between 2019 and 2023, 212 MC twin pregnancies were included in the study. Of these, 24 twin pairs were not delivered in our center, and a further 8 pairs had to be excluded due to perinatal mortality (3 intrauterine fetal deaths and 5 neonatal deaths, precluding within-pair assessment). Following birth, two additional twin pairs were excluded due to genetic or congenital diagnoses (right ventricular outflow tract obstruction and homozygous sickle cell disease). A further 13 twin pairs were lost to follow-up due to relocation abroad or logistical and personal reasons. Of the 164 twin pairs, 83 had umbilical cord blood available for analysis. Missing samples were primarily due to logistical constraints, difficulties in obtaining sufficient cord blood volume, and clinical priorities. Of those, 16 twin pairs were excluded due to consent withdrawal, maternal contamination, incomplete data, and failed DNAm sample-level QC. Ultimately, we studied 67 MC twin pairs comprising of 134 neonates (Figure 1A).

**Figure 1:**
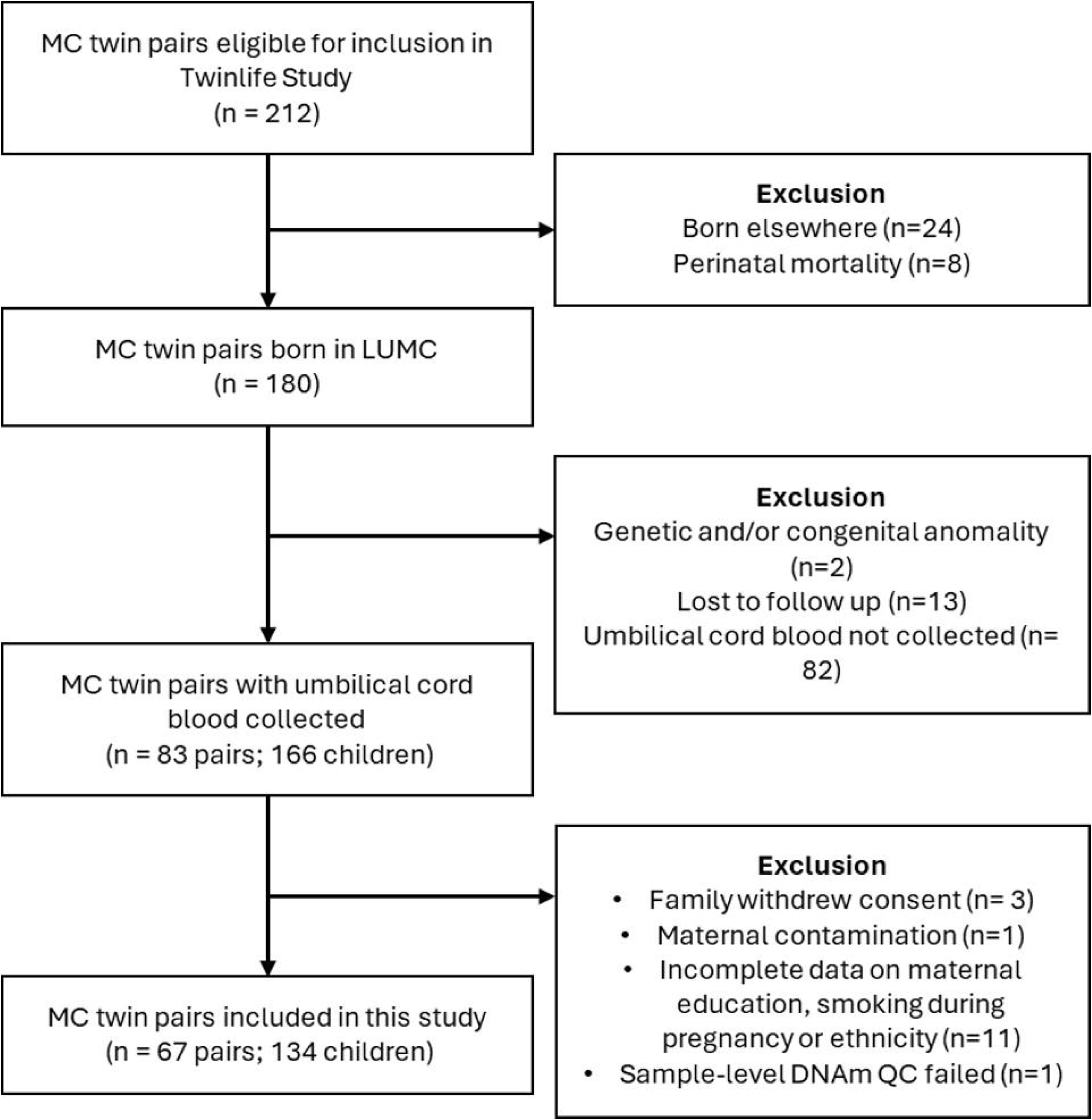
Flowchart of study inclusion. MC = monochorionic, LUMC = Leiden University Medical Center. n= pairs unless otherwise mentioned, DNAm QC = DNA methylation quality control

### Birthweight and covariates

Birthweight was measured in grams immediately after birth. Information on gestational age (weeks), neonatal sex, pre-pregnancy body mass index (BMI), mode of delivery, maternal age at delivery, and parity was obtained from clinical medical records.. Neonates with birthweight <10^th^ centile for gestational age were defined as small for gestational age (SGA) ^31^. Birthweight difference was calculated as birthweight of small co-twin subtracted from birthweight of large co-twin. Birthweight discordance is calculated as birthweight difference relative to birthweight of the large co-twin. Placental share discordance is calculated as the placental share difference between large and small co-twin relative to the placental share of the large co-twin. The twins from a pregnancy were diagnosed with either TTTS (Twin Transfusion Syndrome), sFGR (Selective Fetal Growth Restriction) or TAPS (Twin anemia polycythemia sequence). Neonatal sex was recorded as male or female, mode of delivery as vaginal birth or caesarean section, and parity as nulliparous or multiparous. Maternal educational status, smoking during pregnancy, and ethnicity were self-reported using standardized parental questionnaires. Educational status was categorized as low, secondary, or high; smoking as never, stopped during pregnancy, or continued smoking; and ethnicity as native or foreign. Sentrix position as a technical batch variable was recorded during DNAm preprocessing. To account for cellular heterogeneity in cord blood, cell-type proportions were estimated from DNAm data using the reference-based deconvolution method described by Gervin et al.,^14^ yielding estimates for CD4+ T cells, CD8+ T cells, monocytes, granulocytes, nucleated red blood cells (nRBCs), natural killer cells, and B cells.

### Collection of umbilical cord blood and isolation of mononuclear cells

Umbilical cord blood was collected in citrate-phosphate-dextrose-adenine (CPDA) tubes and processed within 48 hours of collection. All processing was performed under sterile conditions in a laminar flow cabinet. Cord blood was filtered through a cell strainer to remove potential clots prior to further processing. An aliquot of the filtered cord blood (20 µL) was analyzed using a Sysmex XP-300 Automated Haematology Analyzer to determine total white blood cell count (WBC).

The remaining blood was centrifuged at 350 × g for 10 minutes at room temperature, after which the plasma was collected and stored at −80°C. The remaining cellular pellet was resuspended in phosphate-buffered saline (PBS) containing 2% albumin and diluted before being layered onto Ficoll in Leucosep tubes. Mononuclear cells (MNCs) were isolated by density-gradient centrifugation. The enriched mononuclear cell fraction was harvested, washed three times with PBS-albumin, and subsequently cryopreserved.

### Isolation of DNA from isolated cord blood mononuclear cells (CB-MNCs)

Genomic DNA was extracted from cord blood mononuclear cells using the Quick-DNA Miniprep Kit (Zymo Research) according to the manufacturer’s instructions. Briefly, MNC pellets were resuspended in PBS, lysed with Proteinase K at 55 °C, and purified using Zymo-Spin IIC-XLR silica columns with sequential wash steps. DNA was eluted in DNA Elution Buffer (10 mM Tris-HCl, pH 8.5, 0.1 mM EDTA) and stored at −20 °C until further use. DNA concentration was measured using the Qubit fluorometer and NanoDrop spectrophotometer. Samples with a 260/280 ratio ≥ 1.8 were considered of sufficient purity for downstream analyses.

### DNA methylation data

Genomic DNA (500 ng per sample) extracted from CB-MNCs was submitted to the Human Genotyping Facility (HuGe-F) of the Genetic Laboratory of the Department of Internal Medicine at the Erasmus Medical Center (Rotterdam, The Netherlands) where DNAm profiles were generated using the Infinium MethylationEPIC v2 array. Samples were randomized across arrays and plates for sex, the order of inclusion and birth, gestational age, and fetal complications using Omixer v1.8.0.^36^ Twin pairs were positioned adjacent on the same array.

Details of the complete preprocessing pipeline, including R code, quality control procedures, and normalization strategy, are provided in the DNAmArray workflow.^37^ Briefly, raw IDAT files were imported into R using the minfi package. Sample-level quality control was carried out with MethylAid.^2^ Samples failing any of four control-probe–based quality metrics or exhibiting a call rate below 95 were excluded, resulting in the removal of 3 samples. Subsequently, data underwent functional normalization as implemented in minfi,^2^ incorporating five principal components derived from the control probes, followed by probe level QC and imputation for principal component analysis (PCA).

After completion of preprocessing and quality control, the unimputed dataset comprised DNAm measurements for 925,484 CpG sites. Several categories of probes were removed prior to analysis, including those overlapping ENCODE Blacklist regions (5,751 CpGs),^2^ probes classified as polymorphic according to Zhou et al. (24,509 CpGs; Mask information, EPICv2), and CpGs mapping to the X (23,851 CpGs), Y (667 CpGs), or mitochondrial (6 CpGs) chromosomes. Following these exclusions, the working dataset consisted of 867,458 CpG sites. EPIC v2 also has replicate probes for some CpGs and the replicate with the first occurrence in the data frame was retained.

### Birthweight Methylation Profile Score (MPS_BW_)

Of the remaining 867,458 CpG sites in the working dataset, we sought to extract the 914 CpGs that were previously found to be associated with birthweight in the PACE meta-EWAS(P_Bonferroni_ < 1.06 x 10^−7^). Of the 914 PACE-identified CpGs, 72 were absent due to array differences (450K vs EPICv2), and a further 7 were excluded during preprocessing, leaving 835 CpGs for analysis. To derive the MPS_BW_, we followed a previously described method, whereby each CpG’s methylation beta value in our dataset was multiplied by its corresponding regression weight from the PACE meta-EWAS,^8^ These weighted values were then summed across all CpGs, and the resulting score (one per individual) was standardized (z-transformed) with a mean of 0 and 1 standard deviation of 1

### Genotype Data and Polygenic scores for Birth (PGS_BW_)

Genomic DNA (200 ng per sample) extracted from mesenchymal stromal cells from umbilical cord (UC-MSCs) was used for genotyping. 62/64 twin pairs (132/134 neonates) with available DNA were genotyped. Because the twins were monozygotic, i.e., genetically identical twins, only one co-twin per pair was genotyped. The co-twin closest to the cervix was selected for genotyping, as this characteristic is considered to be random. Samples were randomized on plates for sex, the order of inclusion at birth, and tissue type using Omixer v1.8.0.^36^ Genotyping and quality control procedures were conducted at the Human Genomics Facility (Rotterdam, The Netherlands) using the Infinium Global Screening Array (Illumina Inc., San Diego, USA).

Genotype imputation was performed using the Helmholtz Munich imputation server.^9^ Input variant call format (VCF) files, were uploaded in unphased format (hg19 build). Pre-phasing was conducted using Eagle, followed by imputation with the Haplotype Reference Consortium (HRC) panel (version r1.1 2026) across all autosomal chromosomes.^29^ Quality control was performed at the server, including checks for strand alignment, allele mismatches, and genotype consistency. Variants with call rate <90% or allele mismatches were excluded. In total, 806 variants were removed during quality control, leaving 537,273 variants for downstream analyses.

Neonate’s genetic predisposition to birthweight was quantified using a Polygenic score (PGS_BW_), which aggregates genome-wide genetic effects derived from genome-wide association studies (GWAS). In its standard form, a PGS_BW_ is calculated as a weighted sum of trait-associated alleles, where weights correspond to GWAS-derived effect sizes.^11^

To construct the birthweight PGS_BW_, we followed established procedures from the Social Science Genetic Association Consortium^4^ and used summary statistics from the largest available GWAS of birthweight.^39^ SNP weights were computed using LDpred2,^32^ which explicitly models linkage disequilibrium (LD) between variants. Briefly, SNPs were harmonized across the GWAS summary statistics, study genotype data, and the HapMap3+ reference panel by retaining the intersection of variants. LD correlation matrices were then computed using the HapMap3 reference data. The LDpred2 “auto” model was applied, which requires specification of SNP heritability (h²_SNP) and the proportion of causal variants (p). SNP heritability was estimated via LD score regression based on the GWAS summary statistics. The PGS_BW_ was standardized (z-transformed) with a mean of 0 and 1 standard deviation of 1.

### Statistical analyses

To investigate the association between MPS_BW_ and birthweight at the individual level, we first performed an unpaired analysis, with a neonate as the unit of analysis. We used ordinary least squares regression to estimate the association between MPS_BW_ and birthweight across the neonates. This association reflects the combined influence of multiple factors, including but not limited to genetic liability,^3,39^ familial environment, maternal or obstetric factors, as well as shared and non-shared in-utero environmental effects.^6,13,28^ We modeled birthweight as exposure on a continuous scale, scaled to 500-gram units, and standardized MPS_BW_ as the outcome, adjusting for covariates and cell type proportions (S1). To test if common genetic variation influenced the association, the PGS_BW_ was included in the individual-level model as an additional covariate.

To account for dependence of neonates within the same twin pair, cluster-robust standard errors were computed using the CR2 variance estimator, and p-values were obtained using Satterthwaite-adjusted degrees of freedom from the *clubsandwich* package.^33^

We next investigated whether the association between MPS_BW_ and birthweight was also observed within twin pairs. We regressed within-pair differences in MPS_BW_ on the within-pair differences in birthweight, adjusting for batch effects and cell type proportions. This within-pair analysis accounts for characteristics shared by co-twins, including genetic, familial, maternal, and obstetric factors ^12,21,28^.

Detailed descriptions and model specification of the individual-level and within-pair analyses are provided in the Supplementary Methods (S1-S3).

### CpG-level analyses

The analyses described above were repeated for each of the CpGs constituting the MPS_BW._ DNAm levels were quantified using β-values, representing the proportion of methylated signal at each CpG site. Concordance of CpG-level effect estimates was assessed using Spearman’s correlation and effect estimates, comparing the individual-level analysis against (i) the PGS_BW_-adjusted analysis and (ii) the within-pair analysis. Further, directional consistency of CpG effect estimates between each of the analyses perfomed in Twinlife and the original PACE meta-EWAS of birthweight was assessed using Fisher’s Exact Test. For each CpG, the sign of the regression coefficient was classified as positive or negative in each analysis. A 2×2 contingency table was then constructed to compare the distribution of positive and negative effect directions across the two studies. Fisher’s Exact Test was applied to test whether the observed concordance or discordance in directions differed from that expected under independence. The odds ratio and corresponding p-value were used to quantify the strength and significance of directional agreement.

All statistical analyses as well as the generation of graphs were performed using R Software version 4.3.2.

## Results

### Population characteristics

We studied 67 MC twin pairs comprising 134 neonates (Figure 1A). 97% of neonates were born preterm (median gestational age 35.7 weeks (IQR: 32.4-36.4) and 47 neonates (35%) were born small for gestational age. The median birthweight of the study population was 2107 grams (IQR: 1650-2524), with a median birthweight difference of 260 grams (IQR: 139 – 469) and a mean birthweight discordance of 11.7% (IQR: 6 – 21.7) within MC twin pairs. Full baseline maternal characteristics, pregnancy complications, and neonatal characteristics are described in Table 1.

**Table 1:** Maternal, obstetric and neonatal characteristics of 134 MC twin pairs in Twinlife. TTTS: Twin Transfusion Syndrome, TAPS: Twin anemia polycythemia sequence, sFGR: Selective fetal growth restriction, SGA: Small for gestation age

| Characteristics | N = 134<sup>1</sup> |
| --- | --- |
| **Maternal** |  |
| Maternal age at delivery [years] | 32.0 (28.0–35.0) |
| Pre-pregnancy BMI [kg/m<sup>2</sup>] | 22.5 (20.6–25.6) |
| Gestational age at delivery [weeks] | 35.7 (32.4–36.4) |
| **Maternal Ethnicity** |  |
| Native | 116 (87%) |
| Foreign | 18 (13%) |
| **Maternal education** |  |
| Low | 4 (3.0%) |
| Secondary | 42 (31%) |
| High | 88 (66%) |
| **Maternal smoking during pregnancy** |  |
| Never | 100 (75%) |
| Yes, but quit during pregnancy | 28 (21%) |
| Yes, still smoking | 6 (4.5%) |
| **Obstetric** |  |
| Placental share discordance (%) | 26 (11.8 – 44.8) |
| **Mode of delivery** |  |
| Vaginal | 59 (44%) |
| Cesarean | 75 (56%) |
| **Parity** |  |
| Nulliparous | 54 (40%) |
| Multiparous | 80 (60%) |
| **Complications** |  |
| TTTS | 54 (40 %) |
| Laser Surgery | 50 (93%) |
| TAPS | 20 (15 %) |
| sFGR | 24 (18 %) |
| **Neonatal** |  |
| Neonate birthweight [grams] | 2108 (1650–2524) |
| Birthweight difference [grams] | 260.0 (139–469) |
| Birthweight discordance (%) | 11.7 (6.0 – 21.7) |
| **Neonate Sex** |  |
| Male | 62 (46%) |
| Female | 72 (54%) |
| **SGA** | 35% |

<sup>1</sup>Median (IQR); n (%)

### Individual level association

We first examined whether the MPS_BW_ based on healthy pregnancies was also associated with birthweight at the individual level in preterm and often small for gestational age neonates. Indeed, a 500-gram increase in birthweight was associated with a 0.26 SD increase in MPS_BW_ (95% CI [0.09-0.42], p<0.005; Fig 2A). To evaluate whether these associations were partly explained by common genetic variants, the model was further adjusted for PGS_BW_. This yielded a virtually identical estimate of a 0.27 SD increase in MPS_BW_ per 500g increase in birthweight (95% CI [0.08-0.45], p = 0.006; Fig 2A).

**Figure 2.**
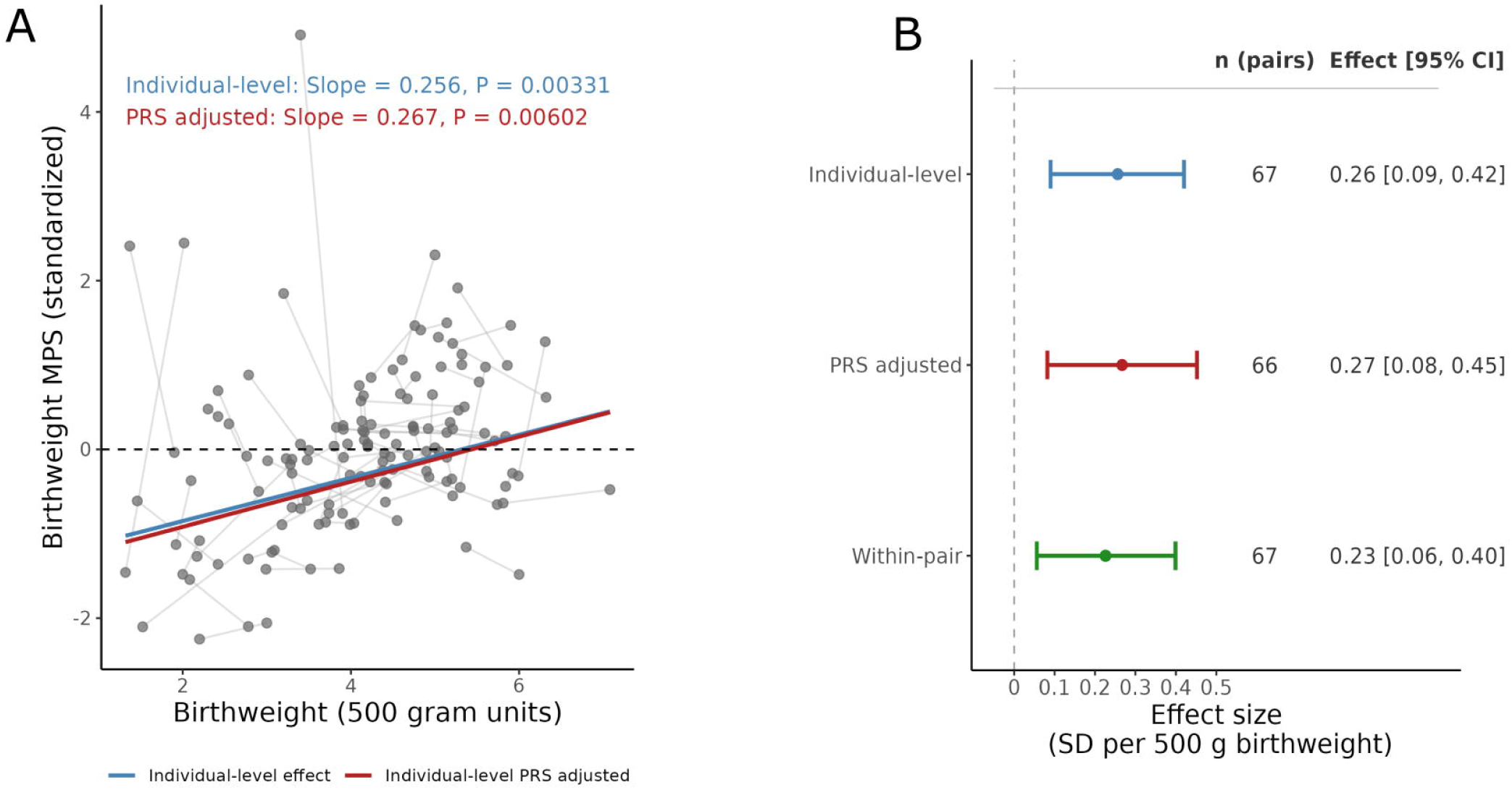
Individual-level, PRS-adjusted, and within-pair association of birthweight with. (A) Scatter plot of birthweight and MPS_BW_, with twin pairs connected by grey lines. The blue and red lines show the fitted linear regression lines from the individual-level and PRS-adjusted analysis respectively. (B) Forest plot of effect estimates from the three analyses, displaying 95% confidence intervals and the number of twin pairs contributing to each analysis; effect sizes are expressed as the change in SD of DNA methylation score per 500 g increase in birthweight.

### Within-pair association

While the individual-level analysis indicates that the MPS_BW_ was associated with birthweight in our population of neonates characterized by preterm birth and SGA, it cannot distinguish whether these associations are induced by unique versus shared (e.g. genetic, familial, maternal and obstetric) influences. Leveraging our twin design, we pesrformed a within-twin pair analysis that controls for potential unmeasured confounders that are shared between the co-twins in a twin pair. For every 500 g increase in the birthweight difference between co-twins, the difference in MPS_BW_ increased by 0.23 SD (95% CI [0.06-0.40], p< 0.05). This was similar to that of the individual-level analysis, in line with the interpretation that the association of the MPS_BW_ with birthweight is not explained by shared genetics, familial environment, or maternal and obstetric factors that are known to influence birthweight (Fig 2B).

### Sensitivity Analyses

We performed sensitivity analyses by repeating the analyses in the expanded sample by excluding covariates with missing data (maternal smoking, maternal education, and ethnicity), thereby including the 11 twin pairs excluded before. This did not appreciably influence effect sizes (Table S1). The same was true for the additional adjustment for pregnancy complications of TTTS and TAPS in the individual-level analysis, suggesting that the respective diagnosis did not affect the association (Table S1).

### Analyses of individual CpGs constituting the MPS_BW_

Next, we assessed whether CpG-level associations showed a similar pattern to that observed for MPS_BW_. We evaluated the consistency of effect estimates for each of the 835 CpGs comprising the MPS_BW_ across the three analyses. CpG-level effect estimates were highly concordant between the individual-level and PGS_BW_-adjusted analyses (slope= 0.97 r_spearman_ = 0.94, p < 0.0001; Fig 3A), suggesting that genetic liability to birthweight did not substantially confound the individual-level results. We next adjusted for all shared effects in the within-pair model and compared effect estimate with the individual-level analyses. We observed a moderate correlation (r_spearman_ = 0.43, p < 0.0001) and a positive association (slope = 0.52, p< .0001), indicating a consistent directional effect but an average attenuation of the effect size in the within-pair model relative to the individual-level model (Fig 3B).

**Figure 3.**
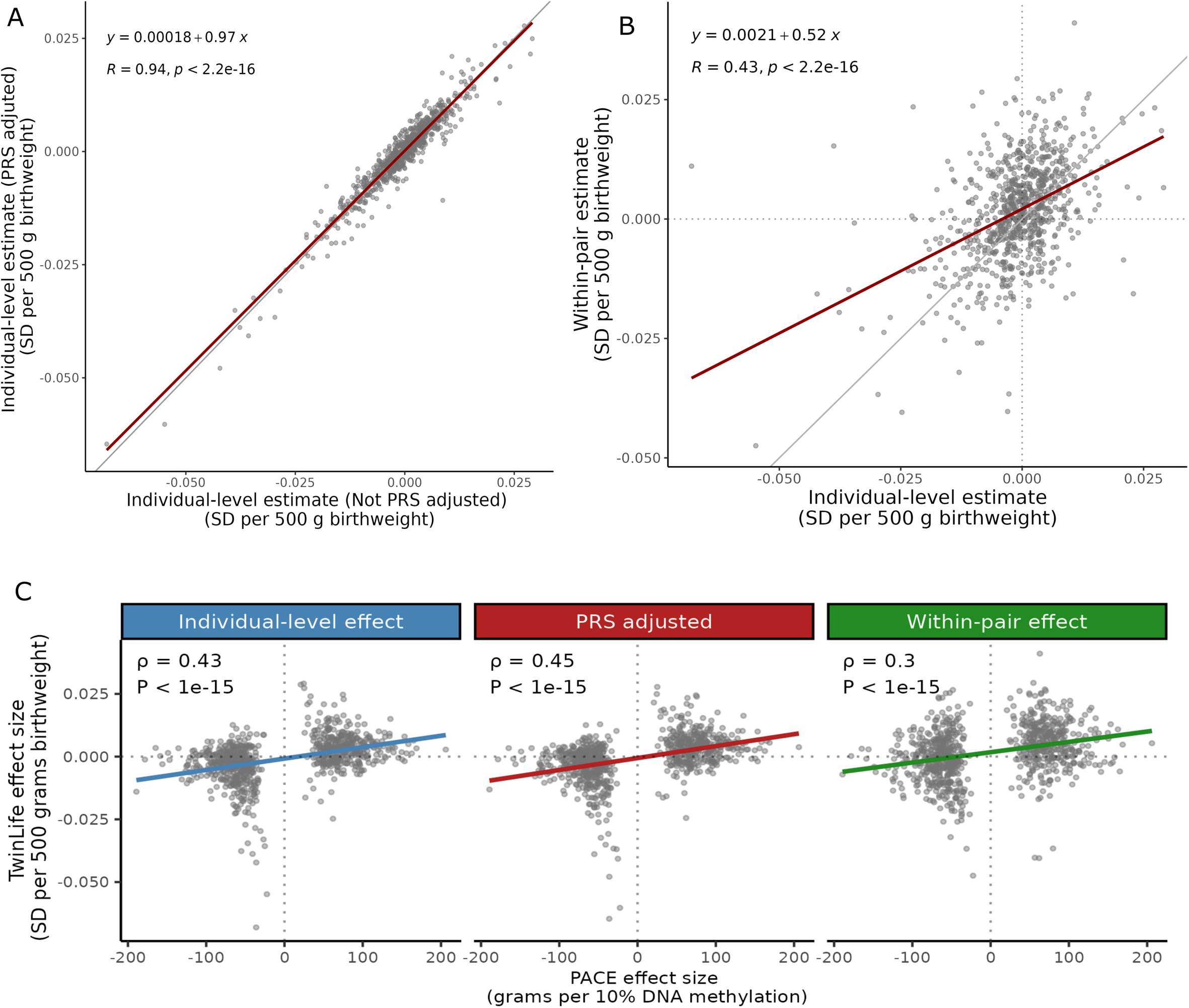
Secondary exploratory analysis of the 835 CpGs constituting the Each dot represents a CpG. (A) Scatter plot of 835 CpGs showing effect estimates from the individual-level model without PGS adjustment (x-axis) versus the individual-level model with PGS adjustment (y-axis); The grey solid line indicates the identity line (***x = y***), and the red line shows the fitted regression line. (B) Scatter plot of 835 CpGs comparing effect estimates of the individual-level analysis with within-pair analysis; Grey dashed lines indicate the x- and y-intercepts, the grey solid line indicates the identity line, and the red line shows the fitted regression line. (C) Faceted scatter plot showing concordance of effect estimates for 835 CpGs between Twinlife and PACE meta-analysis findings; the x-axis shows the PACE effect size (change in grams per 10% increase in DNA methylation), and the y-axis shows the Twinlife effect size (change in standard deviation of MPS_BW_ per 500 g increase in birthweight); grey dashed lines indicate the x- and y-intercepts, and blue, red, and green lines show the fitted slopes from the individual-level, PGS-adjusted, and within-pair analyses, respectively. PGS, polygenic score.

We next explored the agreement between the directionality of effect sizes in Twinlife and that of the PACE meta-analysis. There was high concordance in direction of effect sizes, namely 73% for the individual-level analysis, 72% for the PGS_BW-_adjusted analysis, and 64% in the within-pair analysis (all p < 10^-15^). Likewise, the effect sizes were correlated with those observed in the PACE meta-analysis (r_spearman_ = 0.43, r_spearman_=0.45, r_spearman_ = 0.29, all p < 10^-15^, for the individual-level, PGS_BW-_adjusted, and within-pair analysis, respectively; Fig 3C; Table S2-S4).

## Discussion

We found that a methylation profile score for birthweight (MPS_BW_) derived from the general population generalizes to a high-risk clinical population. This extends previous evidence of associations between DNA methylation and birthweight, which has predominantly been derived from term singleton populations.^25^ Although part of the association at the individual-level analysis may be attributable to direct and indirect genetic influences^8^, we provide evidence using MC twin pairs, that the association is not primarily explained by genetic liability or other factors shared by co-twins, including familial, maternal, and obstetric influences.

Our study leverages the MC twin design to reduce confounding by genetic, familial, maternal, and obstetric factors, allowing us to examine whether within-pair differences in MPS_BW_ are associated with within-pair differences in birthweight beyond these shared influences. In MC twins, unequal placental sharing and vascular connections create a differential intrauterine environment, resulting in unequal nutrient access that can contribute to both birthweight and DNAm differences between co-twins. Thus, the non-shared intrauterine environment provides a plausible source of variation underlying the within-pair association.

The observed association between MPS_BW_ and birthweight may have several interpretations. The MPS_BW_ may mark individual non-shared intrauterine environmental factors that contribute to birthweight differences or DNAm may actually play a role as mediator of such intrauterine influences on birthweight. Alternatively, the MPS_BW_ may be a consequence of birthweight for example by reflecting downstream biological effects of birthweight differences. A similar interpretation has been proposed for epigenetic scores of body mass index (BMI), which have been shown to predict BMI and obesity-related metabolic and clinical outcomes. These scores are often understood as capturing downstream biological embedding of adiposity,^22^ for example through feedback control,^10^ Given that the MPS_BW_ is composed of multiple CpGs, it is possible that different CpGs capture different modalities, and as a composite measure the MPS_BW_ may integrate signals arising from several of these mechanisms (proxy, mediation, or consequence) simultaneously.

In the exploratory analysis of individual CpGs that constitute the MPS_BW_, we found a heterogenous pattern. While correlation was extremely high in the comparison of individual-level analysis with and without PGS_BW_ adjustment, supporting that associations are unlikely to be driven by common genetic liability to birthweight, they were only modestly correlated with the within-pair analysis. This indicated that some CpGs were more sensitive than the others, suggesting that some of the CpGs may reflect shared factors such as familial, maternal, obstetric, or prenatal factors. Overall, however, there was directional concordance with PACE findings across the three analyses, supporting that most of the PACE findings are independent of shared factors.

The use of an MPS_BW_ is a particular strength of our study as it extends its application to a clinically high-risk population of preterm neonates. Since MPS_BW_ provides greater statistical power than single-site approaches like epigenome-wide association analyses,^34^ it offers an advantage in twin studies or clinically relevant populations where sample sizes are inherently limited. In this study, applying an MPS_BW_ enabled targeted validation of the birthweight-methylation signature in a preterm population of neonates.

Our results show that MPS_BW_ is associated with birthweight in preterm neonates often born small for gestational age, and that this association persists after accounting for genetic and other factors shared between co-twins. Although the mechanisms underlying this association remain uncertain, our findings indicate that MPS_BW_ can provide additional predictive information for subsequent health outcomes whether it is in the general population or clinical settings.

## Data Availability

Source data for the main figures in the manuscript with statistical analyses are provided in Supplementary Data file 1. The Twinlife study data underlying this article are not publicly available due to privacy considerations. They can be requested through contact with B.T.H. and shared for replication purposes if replication is conducted within the secure Leiden University Medical Center network environment. Timelines involved in securing access to data vary according to the complexity of the request.

## Acknowledgments

This study was funded by The Dutch Heart Foundation (2017T075) and The LUMC Foundation (BS191 and 123559). We wish to thank the parents and their twins participating in the Twinlife study for their time and effort. This project has received funding from the European Research Council (TEMPO; grant agreement No 101039672 [CC, AN, MS]) and the European Union’s Horizon Europe Research and Innovation Programme (FAMILY, grant agreement 101057529 [CC, AN]). Views and opinions expressed are, however, those of the author(s) only and do not necessarily reflect those of the European Union or the European Health and Digital Executive Agency. Neither the European Union nor the granting authority can be held responsible for them.

## Conflict of interest statement

The authors declare no competing interest.

## Declaration of generative AI and AI-assisted technologies in the manuscript preparation process

During the preparation of this work, the author(s) used "Claude (Anthropic)" and "ChatGPT (OpenAI)" to assist with improving language clarity and grammar of the manuscript. The author(s) reviewed and edited the output as needed and take full responsibility for the content of the published article.

## Supplemental Information

Document S1. Supplemental methods and Table S1

Table S2. Summary statistics of Individual level analysis of 835 CpGs

Table S3. Summary statistics of Within pair analysis of 835 CpGs

Table S4. Summary statistics of Individual level analysis adjusted for Polygenic score of 835 CpGs

## Notes

### Competing Interest Statement

The authors have declared no competing interest.

### Author Declarations

Ethics committee of the Leiden University Medical Center gave ethical approval for this work.

