## Supplemental methods for "DNA methylation signature of birthweight generalizes to high-risk pregnancies and is independent of genetic, maternal, and obstetric factors: a twin study"

### Supplementary Methods:

**S1. Individual level model:** The association between birthweight and MPS was first estimated in a standard multivariable linear regression model, without adjustment for twin pair, to provide a comparison estimate that does not account for shared familial confounding. The model can be expressed as:

$zMPS=\beta_{0}+\beta_{1}*BW_{500g}+\beta_{2}*gestational age+\beta_{3}*parity+\beta_{4} maternal smoking+\beta_{5}*maternal education+\beta_{6}*ethnicit$y + $\beta_{7}* mode of delivery+\beta_{8}*maternal age+\beta_{9}*sex$ + $\beta_{10}*Sentrix_{position}+ \beta_{11}* CD8T+\beta_{12}*NK+\beta_{13}*Bcell+\beta_{14}*Mono+\beta_{15}*Gran+\beta_{16}*nRBCs+ \varepsilon$

In this model, all covariates are entered at the individual level, including gestational age at birth, parity, maternal smoking, maternal educational status, ethnicity, mode of delivery, maternal age at delivery, and neonate sex as pair-level confounders, alongside array position and estimated immune cell type proportions as individual-level technical covariates. Critically, because twin pair is not included, this model the estimated effect of birthweight reflects both within-pair and between-pair associations, and remains susceptible to confounding by all shared familial factors (genetic or environmental). Comparison of β1 from this model with the corresponding within-pair estimate from the twin fixed-effects model allows assessment of the degree to which the association is attributable to shared familial confounding.

**S2. Within pair model:**

This exploits within-pair differences in birthweight to estimate its effect on MPS_BW_ difference independent of shared confounding. The model can be expressed as:

$$zMPS=\beta_{0}+\beta_{1}*BW500g+\beta_{2}*I_{large/small}+\beta_{3}*Sentrixposition+\beta_{4}*CD8T+\beta_{5}*NK+\beta_{6}*B+\beta_{7}*Mono+\beta_{8}*Gran+\beta_{9}*nRBC+\beta_{10.. . 76}*TwinpairIdentifier+\varepsilon$$

With a twin data structure, this approach is mathematically equivalent to the within-twin mean-differencing estimator and ensures that the estimated effects of within-family predictors reflect only variation occurring *between* co-twins within the same twin pair, independent of all factors constant within a twin pair.

To account for shared confounding, twin pair identifier was included as a fixed effect in all models. Specifically, by including a dummy variable for each twin pair identifier, the model absorbs all between-pair variance, such as shared genetic predispositions, familial factors and identifies regression coefficients solely from within-pair contrasts. Unlike the mean-differencing estimator, the dummy variable approach retains one observation per twin, allowing individual-level covariates such as estimated cell type proportions and array position to be included directly as adjustors.

The model was also adjusted for term `I`, which was encoded as 1 for large twin and 0 for small twin. The beta coefficient of `I` quantifies the difference in MPS_BW_ when the difference in birthweight is 0.

**S3. Sensitivity Analysis:**

Individual level and within pair analyses of birthweight and MPS_BW_ in the expanded sample include all samples (n=78 twin pairs) that were removed due to missing data for maternal education, smoking during pregnancy and ethnicity, hence the model did not include these three covariates.

Sensitivity analysis for complications was only done in individual level analysis since co-twins in a twin pair have the same diagnosis label. The individual level analysis was adjusted for the diagnosis of TTTS or TAPS.

**Table S1**

| **Analysis type** | **effect estimate** | **standard error** | **Lower 95% CI** | **Upper 95% CI** | **p-value** |
| --- | --- | --- | --- | --- | --- |
| Expanded sample (individual level) | 0.248 | 1.148 | 0.036 | 0.371 | 0.0017 |
| Expanded sample(Within pair) | 0.204 | 0.084 | -0.016 | 0.422 | 0.0177 |
| Complications adjusted model (individual level) | 0.246 | 0.07 | 0.095 | 0.397S | 0.0022 |
